# Increasing prevalence of bacteriocin carriage in a six-year hospital cohort of *E. faecium*

**DOI:** 10.1101/2024.07.17.24310592

**Authors:** Andrea Garretto, Suzanne Dawid, Robert Woods

## Abstract

Vancomycin resistant enterococci (VRE) are important pathogens in hospitalized patients, however, the factors involved in VRE colonization of hospitalized patients are not well characterized. Bacteriocins provide a competitive advantage to enterococci in experimental models of colonization, but little is known about bacteriocin content in samples derived from humans and even less is known about their dynamics in the clinical setting. To identify bacteriocins which may be relevant in the transmission of VRE, we present a systematic analysis of bacteriocin content in the genomes of 2,428 patient derived *E. faecium* isolates collected over a six-year period from a single hospital system. We used computational methods to broadly search for bacteriocin structural genes and a functional assay to look for phenotypes consistent with bacteriocin expression. We identified homology to 15 different bacteriocins with two having high presence in this clinical cohort. Bacteriocin 43 (bac43) was found in a total of 58% of isolates, increasing from 8% to 91% presence over the six-year collection period. There was little genetic variation in the bac43 structural or immunity genes across isolates. The enterocin A structural gene was found in 98% of isolates but only 0.3% of isolates had an intact enterocin A gene cluster and displayed a bacteriocin producing phenotype. This study presents a wide survey of bacteriocins from hospital isolates and identified bac43 as highly conserved, increasing in prevalence, and phenotypically functional. This makes bac43 an interesting target for future investigation for a potential role in *E. faecium* transmission.

**Importance:** While enterococci are a normal inhabitant of the human gut, vancomycin-resistant *E. faecalis* and *E. faecium* are urgent public health threats responsible for hospital associated infections. Bacteriocins are ribosomally synthesized antimicrobial proteins and are commonly used by bacteria to provide a competitive advantage in polymicrobial environments. Bacteriocins have the potential be used by *E. faecium* to invade and dominate the human gut leading to a greater propensity for transmission. In this work, we explore bacteriocin content in a defined clinically derived population of *E. faecium* using both genetic and phenotypic studies. We show that one highly active bacteriocin is increasing in prevalence over time and demonstrates great potential relevance to *E. faecium* transmission.

## Introduction

Vancomycin-resistant enterococci (VRE) are healthcare-associated pathogens that are transmitted on surfaces and between patients and healthcare workers, resulting in an estimated 54,500 infections and 5,400 deaths each year(1). Most clinical infections with VRE are caused by either *E. faecalis* of *E. faecium*, with vancomycin-resistant *E. faecium* being distinguished from other common nosocomial pathogens by its increased propensity to transmit in hospital settings(2). VRE is endemic in many health care settings resulting in opportunities for patients to acquire VRE during their hospital stay(3, 4). When VRE transmits to a previously uncolonized patient it can, in some patients, dominate in the gastrointestinal community before causing infections(5). The mechanisms that promote this colonization and gut domination by VRE, however, are unclear prompting a need to better understand the underlying biology.

Bacteriocins are ribosomally synthesized antimicrobial proteins and are commonly used by bacteria to provide a competitive advantage in polymicrobial environments(6, 7) . Bacteriocins are produced by many lactic acid bacteria, including enterococci(8) and their production has been suggested to play a role in transmission and intestinal domination, but this role is almost entirely unexplored in the clinical setting. Within the gut, enterococci compete extensively among themselves and with other species(9). In a mouse model of enterococcal gut colonization, it was demonstrated that *E. faecalis* harboring bacteriocin 21 (bac-21) was able to outcompete *E. faecalis* strains lacking this bacteriocin(10). The bacteriocin-producing *E. faecalis* were also able to maintain long-term gut colonization in contrast to the non-bacteriocin producing strain. Gut colonization is a necessary first step in gut dominance(5, 11), and patients colonized with high densities of VRE have a greater amount of shedding into the environment leading to more opportunities for transmission(12). Therefore, bacteriocin production has the potential to increase gut dominance and lead to greater transmission yet an understanding of the specific bacteriocins that could play this role is lacking.

Here, we present genomic analysis of a cohort of 2,428 *E. faecium* blood and perirectal isolates collected over a six-year period from a single hospital system. This sizeable cohort allows for an expansive search for a variety of potential bacteriocin loci, an analysis of how bacteriocin carriage changes over time, and the ability to shed light on how these bacteriocins could promote *E. faecium* transmission in this important setting. We identified many previously described bacteriocins in the patient derived isolates, most of which were found in limited numbers or did not have consistent phenotypic evidence of activity. However, bac43, a plasmid encoded bacteriocin, was found in 58% of isolates and became more prevalent over the study period. Isolates encoding bac43 had phenotypic evidence of bacterial inhibition and were found throughout the phylogenetic tree with evidence of both horizontal and vertical acquisition events.

## Methods

### Ethics

This study was approved by the University of Michigan Institutional Review Board (ID no. HUM00102282), which determined that informed consent was not required as all data utilized were collected for patient treatment purposes.

### Sample collection

*E. faecium* samples were collected from the Michigan Medicine hospital system between 2016 and 2021. Perirectal (PR) swabs were collected as part of the VRE surveillance screening protocol. Patients were screened upon admission to any of the hospital’s nine high risk units and then weekly during their stay in those units. Blood (BL) samples were collected from all patients with *E. faecium* bloodstream infection.

Per hospital protocol, perirectal (PR) swabs were streaked on VRE select plates (Bio-Rad) by the clinical microbiology laboratory. Each positive VRE select plate was transferred to our research laboratory where an isolated pink colony was selected at random, re-streaked twice on Brain Heart Infusion (BHI) agar (Becton, Dickinson and Company, 211065), and stored in BHI broth (Becton, Dickinson and Company, 237500) supplemented with 20% glycerol at -80°C.

Blood (BL) samples were obtained from the clinical microbiology laboratory having been streaked on either BBL TSA II 5% SB or BBL chocolate II agar plates. Random isolated colonies were selected and re-streaked twice on BHI plates with overnight growth at 35°C. If there were multiple clearly distinct morphologies, one of each was selected. Freezer stocks were created from a single colony stored in BHI broth supplemented with 20% glycerol at -80°C.

### Sequencing and assembly

DNA extraction from clones was performed using the Omega Bio-tek Mag-BIND Bacterial DNA 96 Kit (M2350). Illumina library preparation was performed using Collibri PCR-free ES DNA Library Prep Kit for Illumina - Invitrogen (CAT# A39123196) and sequencing was performed using Illumina Nova Seq or HiSeq platform. Reads were processed using a snakemake pipeline for read trimming using Trimmomatic (v0.39)(13), *de novo* assembly using Unicycler (v0.4.8)(14), and annotation using Prokka (v1.14.6)(15). Multilocus sequence types were identified using the software mlst (v2.22.0)(16).

### Hybrid assemblies

Select genomes were assembled using a combination of long and short read data. Long read sequencing was performed using a MinION with a Flongle flow cell from Oxford Nanopore Technologies. Libraries were prepared using the Rapid Barcoding Sequencing Kit (SQK-RBK004) and the Flongle flow cells were primed using the Custom Kit (CUST-KIT). Sequencing data were obtained using the MinKNOW software from Oxford Nanopore Technologies. Adapters were trimmed from long-read sequences using Porechop(v0.2.3)(17), *de novo* assemblies were generated using Unicycler(v0.4.8)(14), and assemblies were annotated using Prokka(v1.14.6)(15).

### Identification of bacteriocins in genome assemblies

A list of bacteriocin protein sequences was obtained from BAGEL4(18), including bacteriocins from classes 1-3. Sequences identical at the amino acid level were removed using the sRNAtoolbox(19). Bacteriocin sequences within our assembled genomes were identified by creating custom BLAST databases of each assembly(20). tblastn was then used to query bacteriocin sequences against each genome assembly database. Output files were filtered using R(v4.3.1). If multiple hits were made to overlapping regions of a genome, the hit with the greatest bitscore was kept. Hits with an e-value greater than 1e-08 and a percent identity less than 60% were removed. Hits from all assemblies were clustered using CD-HIT(21) based on 90% sequence identity and 75% minimal alignment coverage for the longer sequence.

Searches were also performed using nucleotide sequences of other genes involved in bacteriocin production and immunity for enterocin A, bac43, enterocin NKR-5-3B (Supplemental Table 1). These genes were used as the query against each of the custom BLAST databases for each genome assembly.

### Variant calling and phylogenetic analysis

Variants were identified using a custom snakemake pipeline. Trimmed reads were mapped to an internal reference strain *E. faecium* BL00198-1 and variants were called using snippy(22). A core genome was produced using snippy-core; samples with less than 250,000 base pairs aligned were excluded. The core alignment was then cleaned using snippy-clean_full_aln, and a phylogenetic tree was produced using Gubbins(23). Gubbins was run for six iterations where the first tree built using ‘rapidnj’ and ‘JC’ model. The remaining five iterations were built using ‘raxmlng’ and the ‘GTR’ model. Samples with less than 75% homology to the reference strain were excluded. The tree figure was rendered and annotated using ggtree in R(v4.3.1). MLSTs with less than 20 isolates associated with them are classified as “Other”. Bacteriocin sequences with less than five isolates associated with them are classified as “Other”.

### Variation analysis of enterocin A gene cluster

Variants in the enterocin A gene cluster and the bacteriocin 43 encoded plasmid were identified using an internal snakemake pipeline. For enterocin A, trimmed reads were mapped to internal reference with the entire enterocin A gene cluster (BL02040-1). Variants were called using FreeBayes(24) with the following parameters: minimum base quality greater than or equal to 10 and minimum alternate fraction less than or equal to 0.5. Variants were annotated using SnpEff(25). Insertion elements were identified and annotated using panISa(26) and ISFinder(27). Areas of low read coverage indicating gene deletions were identified using SAMtools(28) depth and filtering to positions with at least 10% of the average coverage across the whole gene locus. Variants, including insertion elements, identified in deleted regions were excluded using custom R scripts (v4.3.1). Read pileups were visualized using igv-reports(29). Variants and low coverage regions were manually inspected using these read maps; mapped regions were identified in isolate genome assemblies to confirm presence or absence of variants and low coverage regions.

### Variation analysis of bac43 encoding plasmid pDT1

Trimmed reads were mapped to an internal reference isolate, PR46485-1-C1, which encodes a plasmid identical to the previously published pDT1 sequence(30). Variant calling, annotation, identification of deletions, and insertion element identification was repeated for pDT1 with the same methodology as for enterocin A. Additional analyses for pDT1 included coverage calculations across the plasmid and bac43 specific genes using SAMtools(28) coverage. Isolates with less than 45% mean depth or less than 55% coverage across the plasmid were excluded from further analysis. Variant positions were accepted if the RO (reference observations) to DP (total reads mapped) ratio was less than 10%, and if the RO was less than 30 reads. Percent identity was then calculated as the number of non-variant positions divided by the total number of bases called. Isolates were finally filtered by those with at least 99.5% identity to the aligned portions of the pDT1 plasmid or if the BLAST analysis identified the full structural gene of bac43.

### Visualizing genetic associate among variants

Genetic associations between variants in the enterocin A gene cluster and pDT1 were visualized using PHYLOViZ(31) (v2.0) using the goeBURST Full MST algorithm (goeBURST distance). Subcluster annotations were made based on shared variants and/or variations within the same genomic regions. Gene homology maps were created using Easyfig(32) (v2.2.2) with a minimum identity threshold of 60%.

### Bacteriocin activity assay

Putative bacteriocin activity was assessed using a modified soft-agar assay for bacteriocin production and immunity(30). Isolates were streaked from freezer stock onto BHI agar plates and incubated at 35°C overnight. Single colonies were resuspended in 50ul saline and poked into a soft agar lawn containing a bacteriocin sensitive strain. If results were discordant with functional predictions, a higher inoculum was tested by using a toothpick to poke a single colony directly into the lawn. After incubation at 35°C overnight, putative killing effect was determined by the presence of a halo of bacterial clearing around the poke into the bacteriocin sensitive lawn. Immunity to enterocin A was determined poking an enterocin A producer (BL02040-1) into a soft agar bacterial lawn of the strain being tested and looking for the absence of a halo of bacterial clearing.

Immunity to bac43 was determined by the ability to grow in the presence of supernatant derived from a bac43 producing strain (PR46485-1-C1). Bac43 supernatant was obtained by incubation of PR46485-1-C1 in 45mL BHI at 35°C, 130rpm, for 24 hours. The culture was centrifuged for 15 minutes at 3220rcf and the supernatant was filter sterilized twice (0.22um filter). Colony resuspensions were plated on BHI agar supplemented with 10% bac43 supernatant. Plates were incubated at 35°C overnight. Immunity to bac43 was determined by bacterial growth on BHI plates supplemented with 10% bac43 lysate.

Isolates were chosen for the bacteriocin production and resistance assays as follows. When sequences were predicted to be nonfunctional (i.e. deletions or insertion elements present in bacteriocin genes) only one isolate with that mutation was tested. If the isolate with the presumed non-functional bacteriocin was found to be negative on the bacteriocin production and immunity assays, other isolates carrying the mutation were not tested. For sequence isolates from multiple MLSTs, one isolate was tested from each of the MLSTs present in at least 20 of all isolates.

### Data availability

All scripts and pipelines are available on GitHub (https://github.com/woodslab/bacteriocin_search). The whole genome sequencing project for our reference *E. faecium* isolates have been deposited at DDBJ/ENA/GenBank: BL00198-1 (accession JBEFKN000000000), BL02040-1 (accession JBEFKM000000000), PR46485-1-C1 (JBEFKL000000000). The versions described in this paper are JBEFKN010000000, JBEFKM010000000, and JBEFKL010000000, respectively.

## Results

### Samples included

The cohort for this study included the first *E. faecium* isolate from each patient with a VRE *faecium* positive rectal surveillance swab between 2016 and 2021. This consisted of 2300 patients (Table 1). Additionally, 278 patients were found to have *E. faecium* growth in blood cultures. The first isolate from surveillance and blood culture per patient was sequenced. One-hundred and twenty-eight patients had both a perirectal (PR) swab and a blood (BL) sample taken during their stay in the hospital resulting in a total of 2428 *E. faecium* isolates.

**Table 1:**
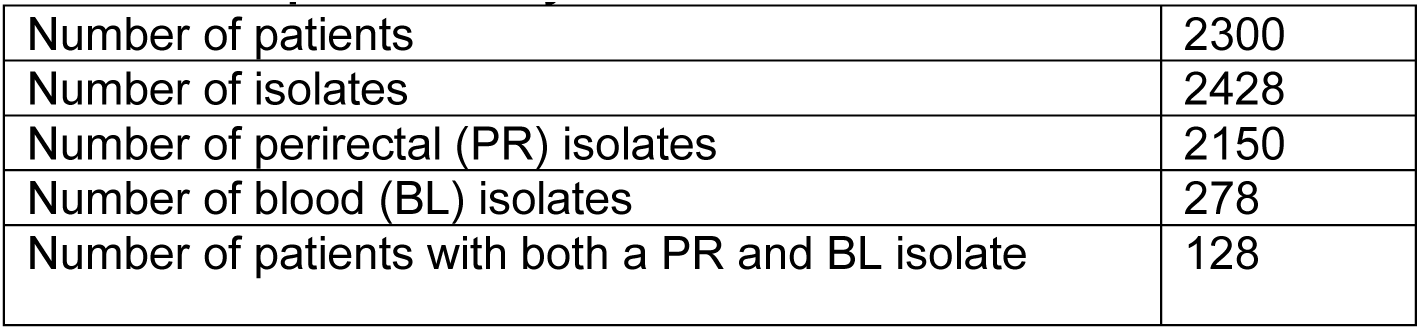
Sample summary.

### Bacteriocins identified in the clinical cohort

BLAST hits to bacteriocin structural genes in our cohort were clustered based on sequence identity. There were 17 bacteriocin clusters identified with homology to 15 different bacteriocins (Figure 1, supplemental table 3). Most bacteriocin sequence clusters were rare, being found in less than 3% of isolates. The majority of these low frequency clusters have previously been characterized from *E. faecium*(33–39), but enterocin SE-K4(40) and bac31(41) were characterized from *E. faecalis* and pneumolancidin(42) was characterized from *S. pneumoniae*. We chose the four most common cluster hits to investigate further as their higher prevalence suggested a potentially relevant role in hospital transmission. This includes the enterocin A cluster (98% of isolates), two cluster hits to bac43 (57.9% of isolates and 40.7% of isolates, respectively), and enterocin NKR-5-3B cluster (19.2% of isolates)(Figure 2A-D).

**Figure 1:**
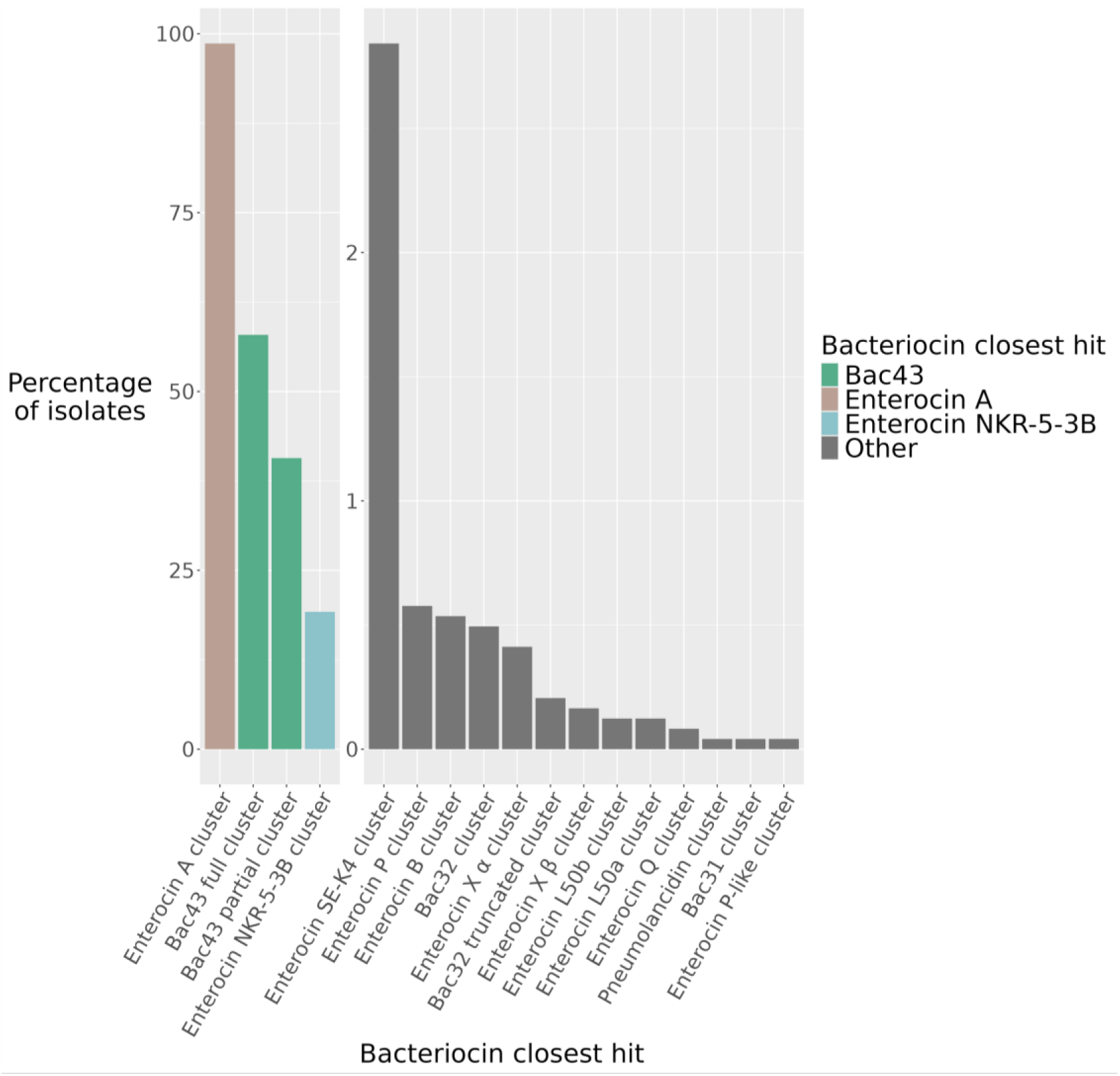
Frequency of bacteriocin gene sequence hits. Bacteriocin sequences were classified into 17 different clusters with homology to 15 different bacteriocins. Four bacteriocin clusters were identified as present in over 19%: enterocin A cluster, bac43 full cluster, bac43 partial cluster, and enterocin NKR-5-3B cluster.

**Figure 2:**
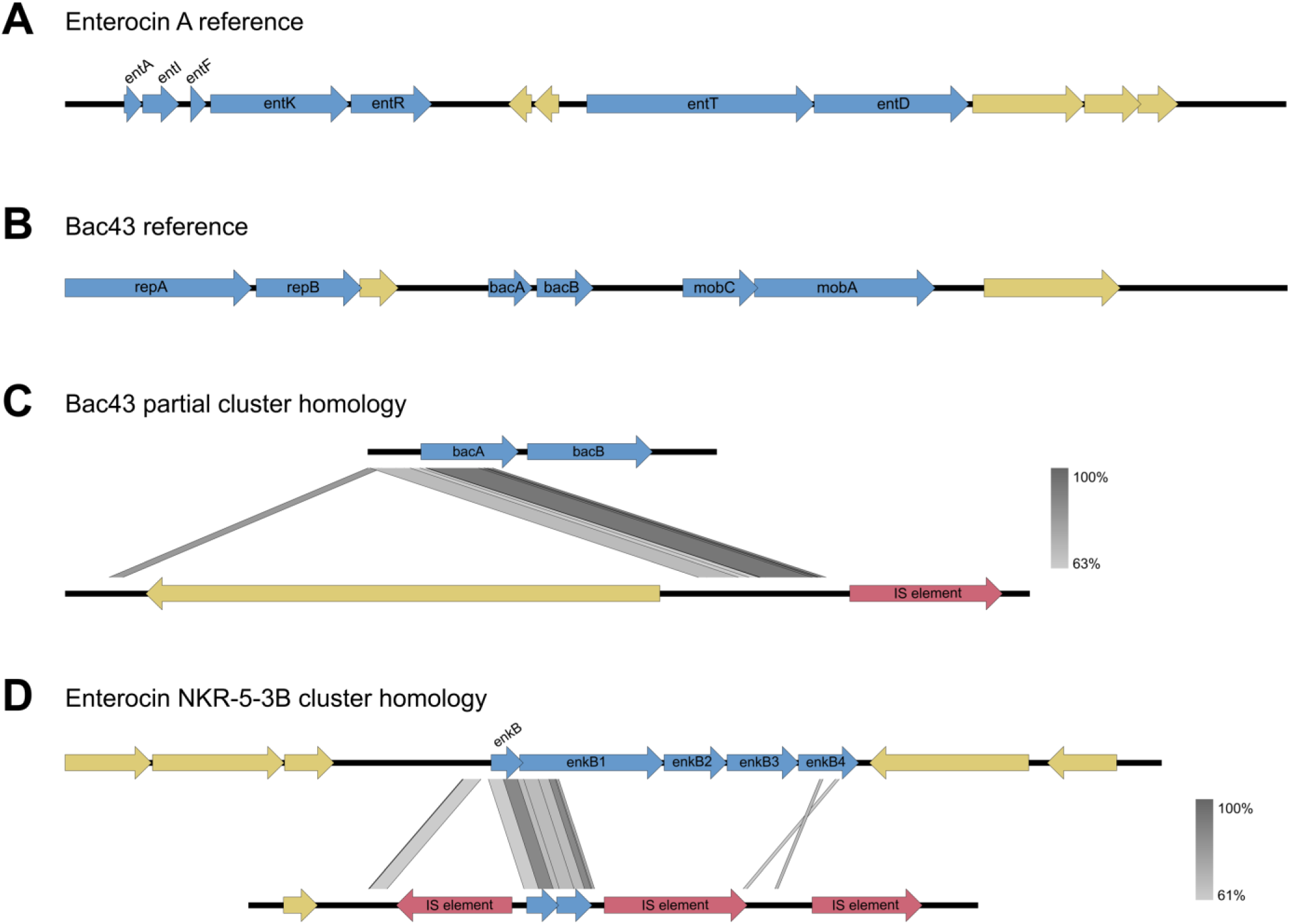
Gene maps and homology to previously identified bacteriocins. Genes related to bacteriocin production are blue, hypothetical proteins are yellow, and insertion elements are red. A. Internal reference of the enterocin A gene cluster. B. Internal reference of the bacteriocin 43 encoding plasmid pDT1. C. Homology of the bacteriocin 43 genes to the bac43 partial cluster BLAST hit. D. Homology of the enterocin NKR-5-3B genes to the enterocin NKR-5-3B cluster BLAST hit.

The enterocin NKR-5-3B cluster shares 87.3% amino acid homology to the structural gene *enkB* of enterocin NKR-5-3B and was identified in 19.2% of isolates. This class IIc circular bacteriocin was previously characterized on a 9kb chromosomal region of *E. faecium*(43). The previously described functional operon includes five genes associated with enterocin NKR-5-3B: *enkB* the bacteriocin structural prepeptide, *enkB1* and *enkB3* for secretion and self-immunity, *enkB2* for maturation, and *enkB4* for self-immunity. None of the clinical isolates contained the entire previously described 9kb region. Rather we found homology to *enkB* and approximately 200 bp of *enkB1* (Figure 2D). Given the lack of predicted function, this cluster was not further investigated.

### Analysis of the enterocin A clusters

The enterocin A structural gene, *entA,* was identified in 98.6% of isolates(44, 45). Enterocin A is a class IIa bacteriocin previously characterized in *E. faecium* strain CTC492. In that strain, the enterocin A gene cluster is encoded on a 10.5kb region of the bacterial chromosome(45) (Figure 2A), and the locus includes genes for structural protein (*entA)*, self-immunity (*entI)*, the peptide pheromone (*entF)*, the two-component regulatory system (*entK* and *entR)*, and the peptide transporter (*entT and entD)*.

We identified variant positions, insertion elements, and deletions by comparing isolates to an internal reference. This reference sequence has a missense variant in the *entI* gene (position 246717 in BL02040-1, G to A) compared to the previously published sequence(45). We identified 87 unique sequences of the enterocin A gene cluster in our collection, 61 of which were found in only a single isolate. These sequences were assigned to five subclusters labeled A through E (Figure 3). Subcluster A is centered around sequence 1, which is found in 84% of all isolates. Sequence 1 isolates have a large deletion spanning *entFKRTD*. Of the 41 other sequences in subcluster A, 29 share this deletion. Four sequences in subcluster A (4, 6, 9, and 12) have all the genes required for enterocin A function intact. In subcluster B, four sequences (3, 49, 73, and 85) also have all the enterocin A genes. The other sequence in subcluster B (35) has an insertion element in histidine protein kinase *entK* likely interrupting function. Subcluster C sequences have an insertion element that is likely inactivating the regulator gene *entR.* Subcluster D shares the insertion element in *entR* with subcluster C plus an additional insertion element the immunity gene *entI* also likely interrupting function. Subcluster E has three sequences which all have the complete enterocin A gene cluster but with approximately 30 synonymous and missense mutations.

**Figure 3:**
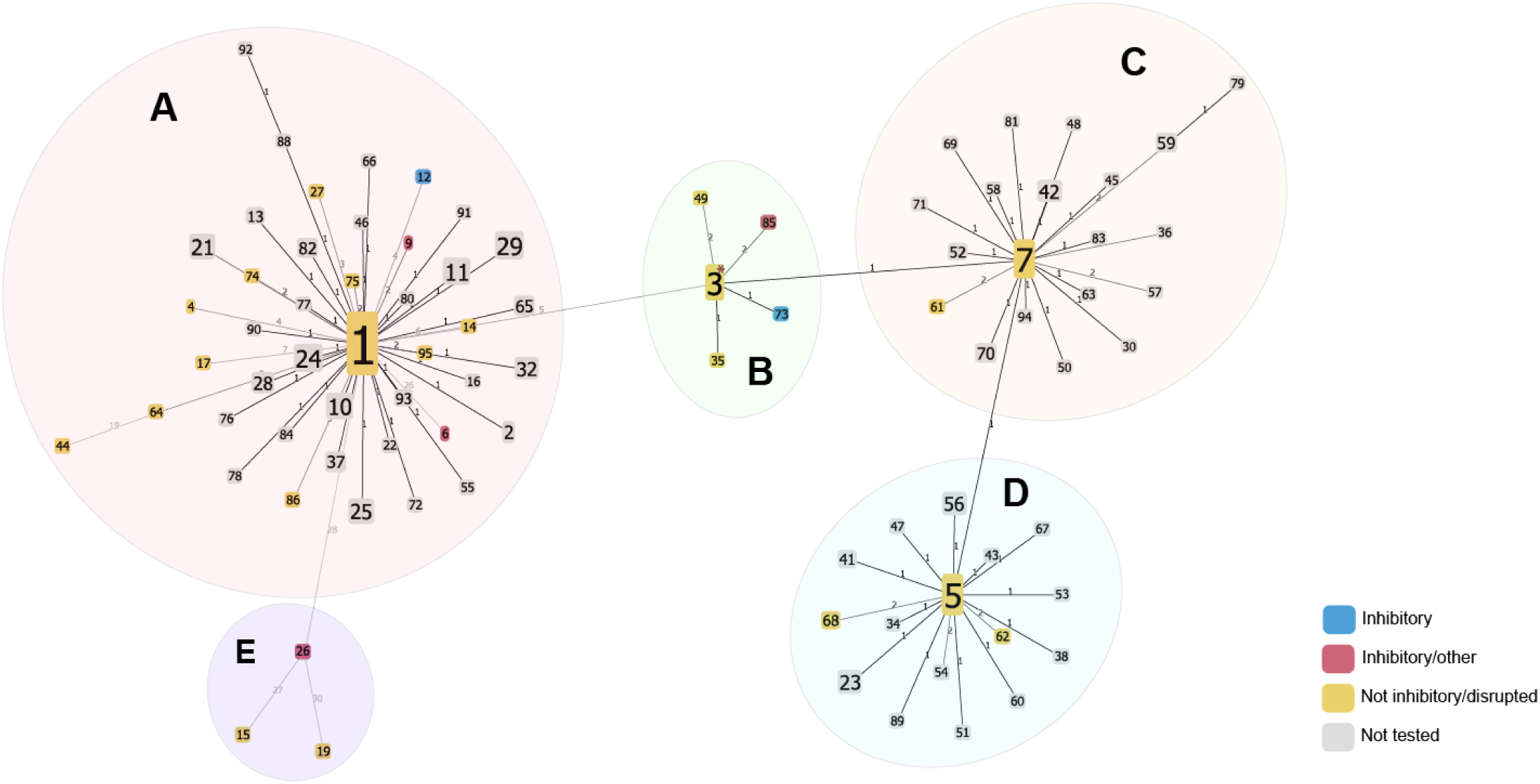
Enterocin A sequence clusters. Isolates were assessed for SNPs, insertion elements, and gene deletions within the enterocin A gene locus and are clustered based on matching patterns of variations (sequences). Each node is a unique sequence of the enterocin A locus and branch numbers represent the number of variations between the sequences. Node size increases based on the number of isolates belonging to that sequence. The results of phenotypic assays on tested strains are noted on each sequence node. Inhibitory sequences (blue and red) have both the full enterocin A locus and can kill a bacteriocin sensitive strain, inhibitory/other sequences (red) also encode other bacteriocins. Not inhibitory/disrupted sequences (yellow) either lacked inhibition in phenotypic assays and/or had disruptions in the enterocin A locus that would predict nonfunction. Sequence 3 isolates are marked as non-inhibitory due to mixed phenotypic results.

### Analysis of the bac43 gene clusters

Bacteriocin 43 (bac43) is a class IIa *sec*-dependent bacteriocin previously isolated from clinical vancomycin resistant *E. faecium* isolates(30). The 6.1kb plasmid pDT1 encodes the bac43 structural gene *bacA*, the gene for self-immunity *bacB*, plasmid mobilization genes *mobC* and *mobA*, and plasmid replication genes *repA* and *repB*(30)(Figure 2B).

Bac43 was identified as the closest bacteriocin hit in two common clusters. One of these, referred to as ‘bac43 partial’ cluster, was found in 40.7% of isolates and aligned only to part of the *bacA* coding region, has no homology to the immunity gene, and the start codon found on *bacA* was absent. Thus, we do not expect it to be functional. The aligned bac43 partial sequence displayed only 87.5% amino acid homology to previously published *bacA*, and were not found on contigs with other pDT1 genes (Figure 2C).

The second cluster, ‘bac43 full,’ was identified in 57.9% of isolates and displayed 99.5–100% amino acid homology to the published *bacA*. Sequences falling into the bac43 full cluster also have homology to the immunity gene *bacB* and are often found on contigs consistent with the pDT1 plasmid.

Variants in the pDT1 plasmid were identified by comparing isolates to an internal reference genome carrying a plasmid identical to the one previously published(30). Four isolates discordant with phenotypic predictions were assayed using PCR for the presence of the *bacA* (Supplemental Table 2). PCR revealed an absence of this gene, and these isolates were removed from further analysis. We identified 106 unique sequences of the bac43 encoding plasmid pDT1, 77 of which were found in only one isolate. These sequences are organized into nine subclusters (Figure 4). Subcluster A is centered around sequence 3 which is found in 39.7% of isolates and is identical to the published pDT1 sequence(30). Most of the variation within this subcluster is outside of *bacA* and *bacB*, but four sequences had a mutation in one of these genes: a synonymous variant in *bacA* (sequence 82), an insertion element in *bacB* (sequence 14), and two with insertion elements in different positions in *bacA* (sequences 57 and 84). Additionally, sequence 65 (subcluster H) has a missense variant in *bacB*. Sequences 9 and 73 (subcluster D) have lost the bac43 genes but retained portions of the pDT1 plasmid. The remainder of the sequences are characterized by insertion elements and/or alterations to the plasmid backbone.

**Figure 4:**
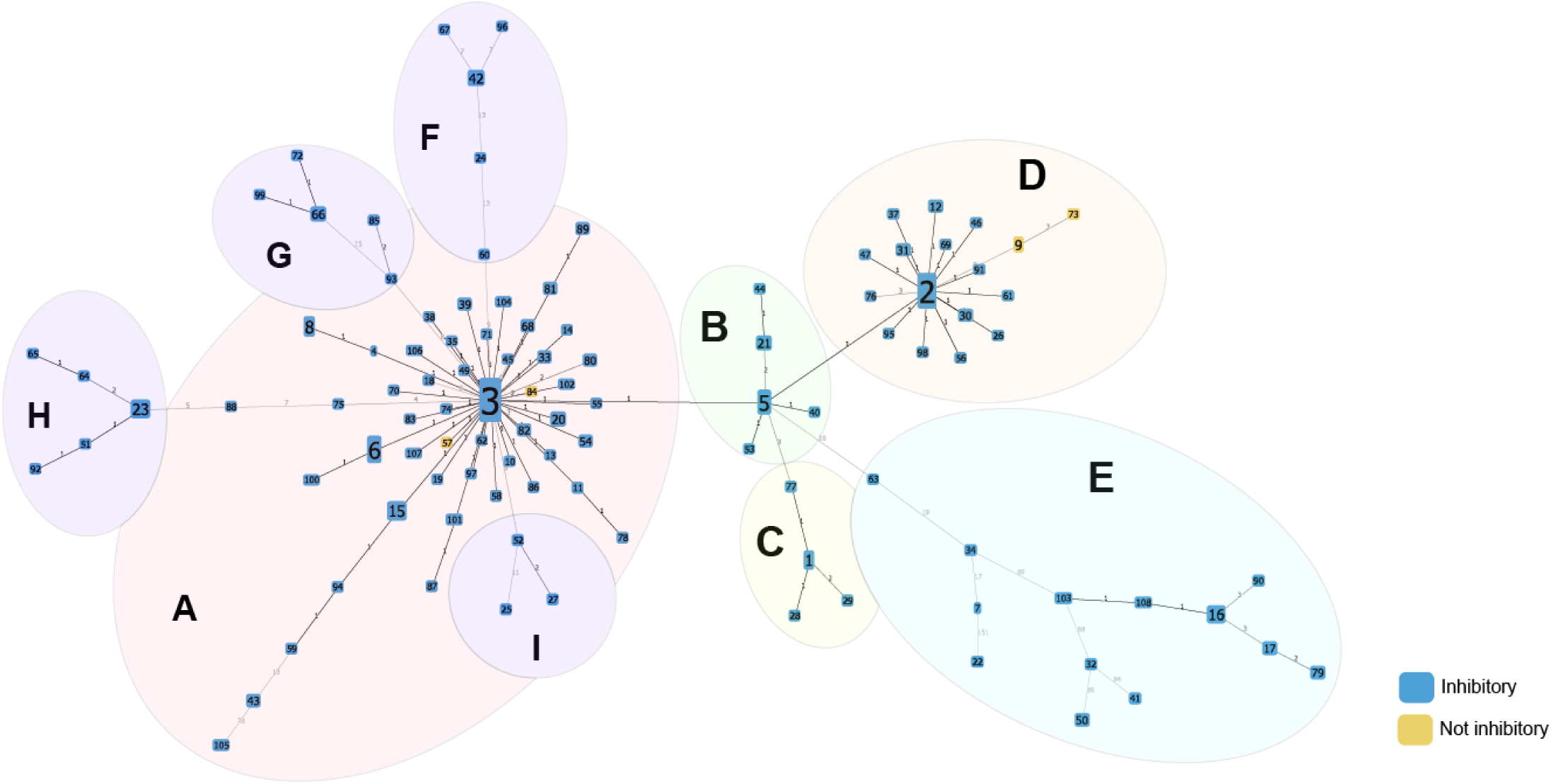
Clustered variations within the bac43 encoding plasmid pDT1. Isolates were assessed for variants, insertion elements, and gene deletions within the bac43 encoding plasmid pDT1 and are clustered based on matching patterns of variations (sequences). Each node is a unique sequence of pDT1 and branch numbers represent the number of variations between the sequences. Node color represents phenotypic testing results. Node size is determined by number of isolates belonging to that sequence. Inhibitory sequences (blue) can kill a bacteriocin sensitive strain.

### Phenotypic evaluation of bacteriocin containing isolates

Phenotypic assays were performed to determine whether strains produce a phenotype of (1) killing of a strain with no bacteriocin sequences and (2) immunity to killing by strains that encode a known bacteriocin producer (for enterocin A) or a crude extract (for bac43).

For strains encoding the enterocin NKR-5-3B-like sequence, we tested seven isolates from different MLST backgrounds. As expected, none of the isolates demonstrated inhibition against a bacteriocin sensitive strain.

For enterocin A, eleven sequences were classified as potentially functional based on the absence of deletions or insertion elements into the gene cluster. Isolates encoding six of these sequences showed both consistent killing of the sensitive strain and resistance to killing by an enterocin A producer consistent with a fully functional enterocin A locus (sequences 6, 9, 12, 26, 73, and 85)(Figure 5A); four of these isolates (sequences 6, 9, 26, and 85), however, also encode other bacteriocins such as enterocin B, enterocin P, enterocin X𝛼, and enterocin SE-K4 which could not be ruled out as the source of inhibition. The five sequence 3 isolates that were tested gave mixed results. Four of the five tested isolates encoding sequence 3 demonstrated an inhibitory and resistant phenotype that could be consistent with a functional enterocin A locus, but the contribution of other bacteriocins could not be excluded. The only sequence 3 isolate that encodes no other bacteriocins had no inhibitory activity but did demonstrate resistance to killing by enterocin A. This pattern of being resistant but noninhibitory was also demonstrated by sequence 49. As neither of these isolates encode other bacteriocins the enterocin A resistance could stem from *entI* expression or chromosomal mutations. The remaining three sequences did not have an inhibitory phenotype. The sequence 4 isolate showed resistance without inhibition but unlike the preceding isolates, the tested isolate also encodes other bacteriocins. Sequence 15 and 19 lack both inhibitory activity and enterocin A resistance; both have numerous mutations that could have resulted in non-function (Figure 5B).

**Figure 5:**
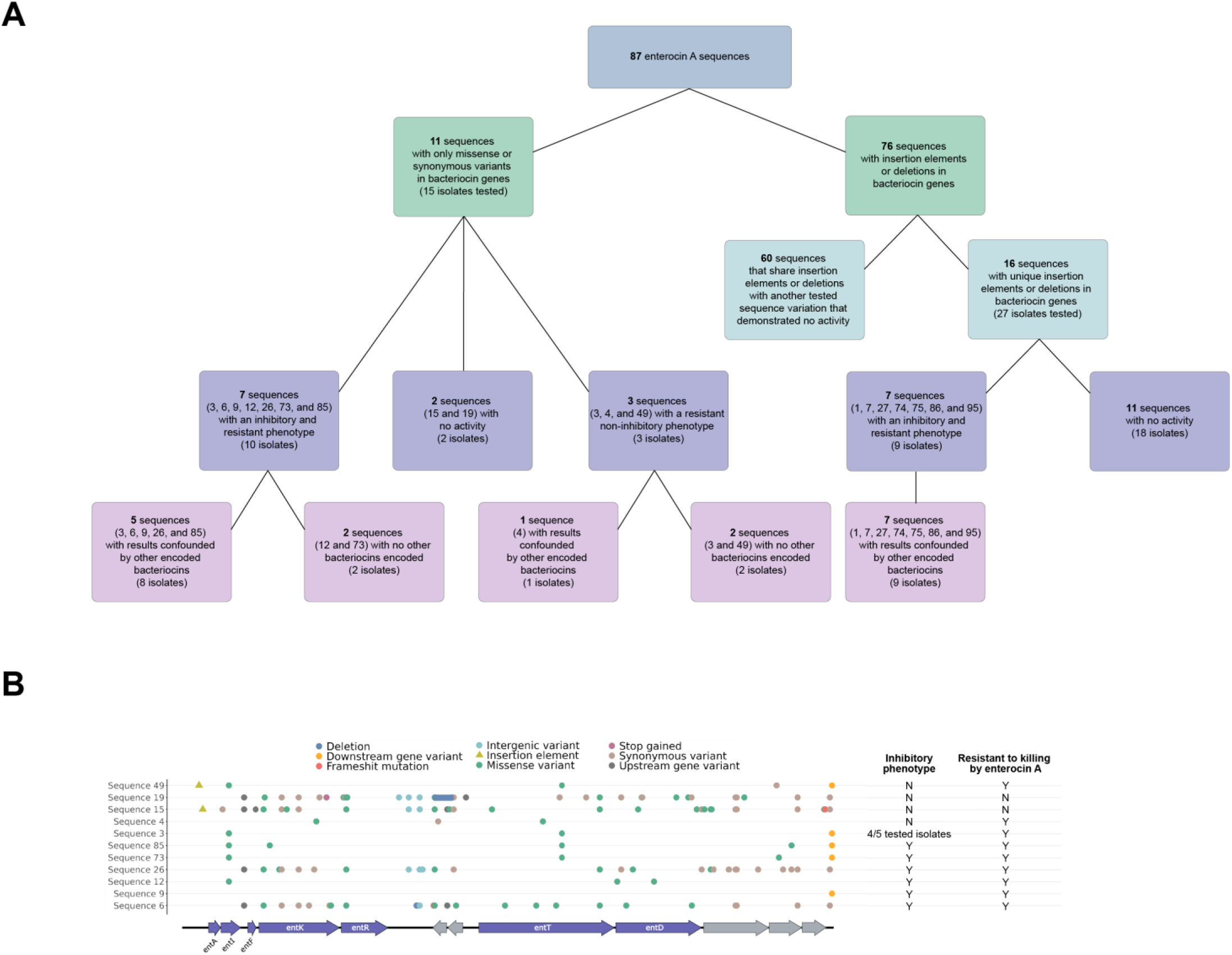
Enterocin A phenotypic analysis. A. Overview of enterocin A isolates tested, functional expectations, and experimental results. B. Gene maps aligned with the enterocin A reference locus for sequences that were expected to be functional (i.e. no deletions or insertion elements within the bacteriocin genes).

Sixteen sequences were predicted to be non-functional due to deletions of or insertions into genes essential for enterocin A production. All of the tested isolates had a phenotype that was neither resistant nor inhibitory in isolates that did not encode other bacteriocins. Seven isolates did encode other bacteriocins and these had both inhibitory activity and grew in the presence of the enterocin A producer, suggesting the other bacteriocin loci, rather than the enterocin A locus, were the cause of the assay results.

Phenotypic testing was performed on 132 isolates representing 106 sequences with pDT1 variants (Figure 6A). Ninety-nine of these had 100% conserved sequences of *bacA* and *bacB* but other mutations in the pDT1 plasmid; all 124 isolates tested with these 99 sequences were able to kill the bac43 sensitive strain and were resistant to killing by a bac43 producer. Among the five sequences with mutations in the bac43 genes, three retained both an inhibitory phenotype and bac43 resistance: sequence 65 (missense variant in *bacB*), 82 (synonymous variant in *bacA*), and 14 (insertion element in *bacB*). However, no inhibition or resistance to bac43 were observed by isolates with an insertion element in *bacA* (sequences 84 and 57). Because the variant analysis aligned sequences to the full pDT1 plasmid, two sequences (9 and 73) were identified that lost the bac43 genes but retained portions of the plasmid (Figure 6B); these isolates did not demonstrate inhibition or resistance to bac43.

**Figure 6:**
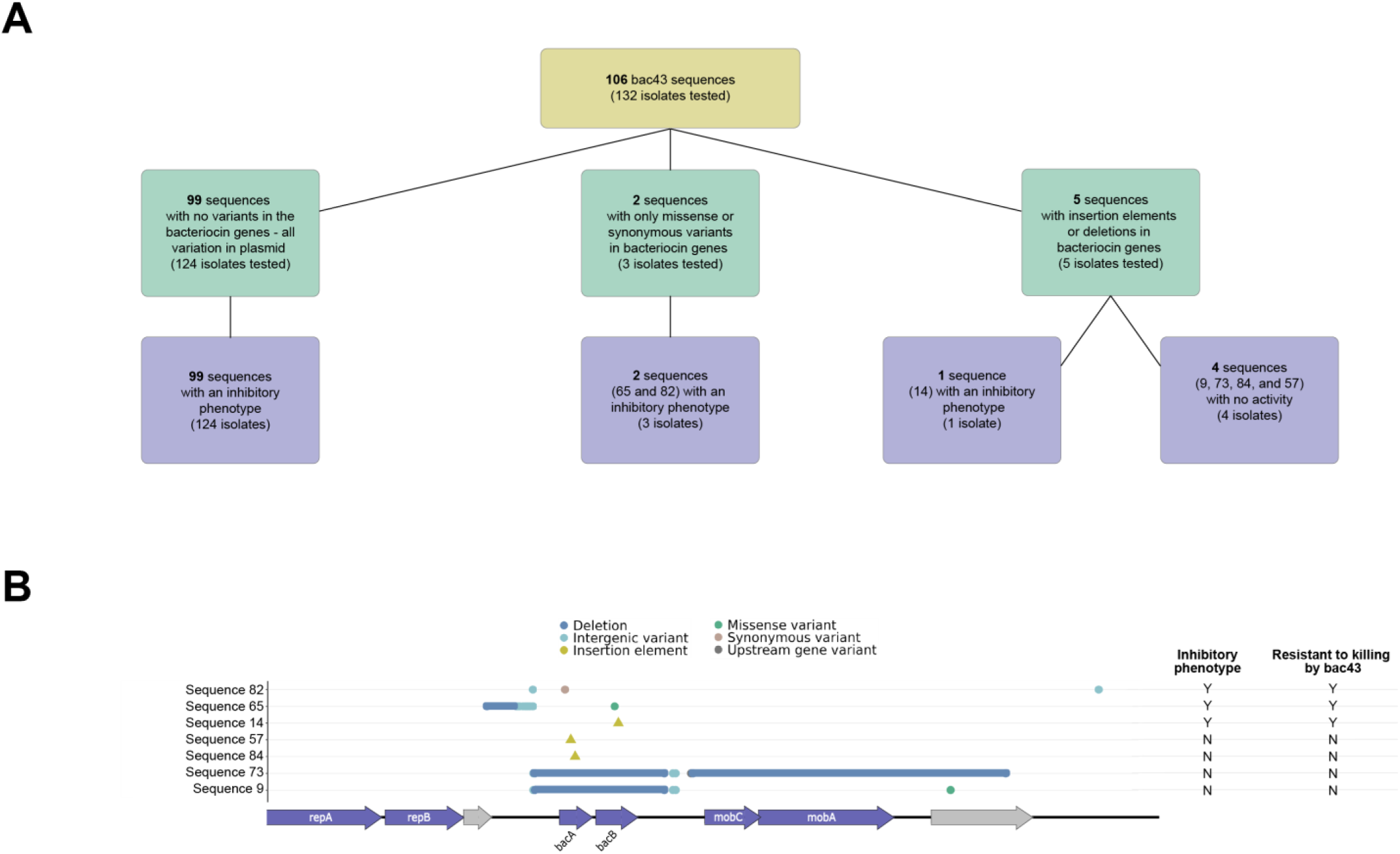
Bac43 phenotypic analysis. A. Overview of bac43 isolates tested, functional expectations, and experimental results. B. Gene maps aligned with the bac43 encoding plasmid pDT1 for sequences with variants in the bac43 genes *bacA* and *bacB*.

### Phylogenetic analysis

A phylogenetic tree of the isolates was created to examine the evolutionary relationships between the bac43 and enterocin A sequences. The enterocin A sequences cluster together supporting a predominantly vertical mode of transmission (Figure 7A). Sequences are not restricted to MLSTs: for example, sequence 1 with a deletion of *entFKRTD* is found in multiple MLST backgrounds suggesting this wide-spread deletion may have occurred before MLST differentiation. Five of the seven phenotypically functional isolates encoding the full enterocin A locus (six sequence variations plus the enterocin A reference) passed the homology thresholds for inclusion on the tree. Four of these isolates cluster together while one is found in another cluster surrounded by nonfunctional sequence 1 isolates consistent with horizontal gene transfer of the intact gene locus.

**Figure 7:**
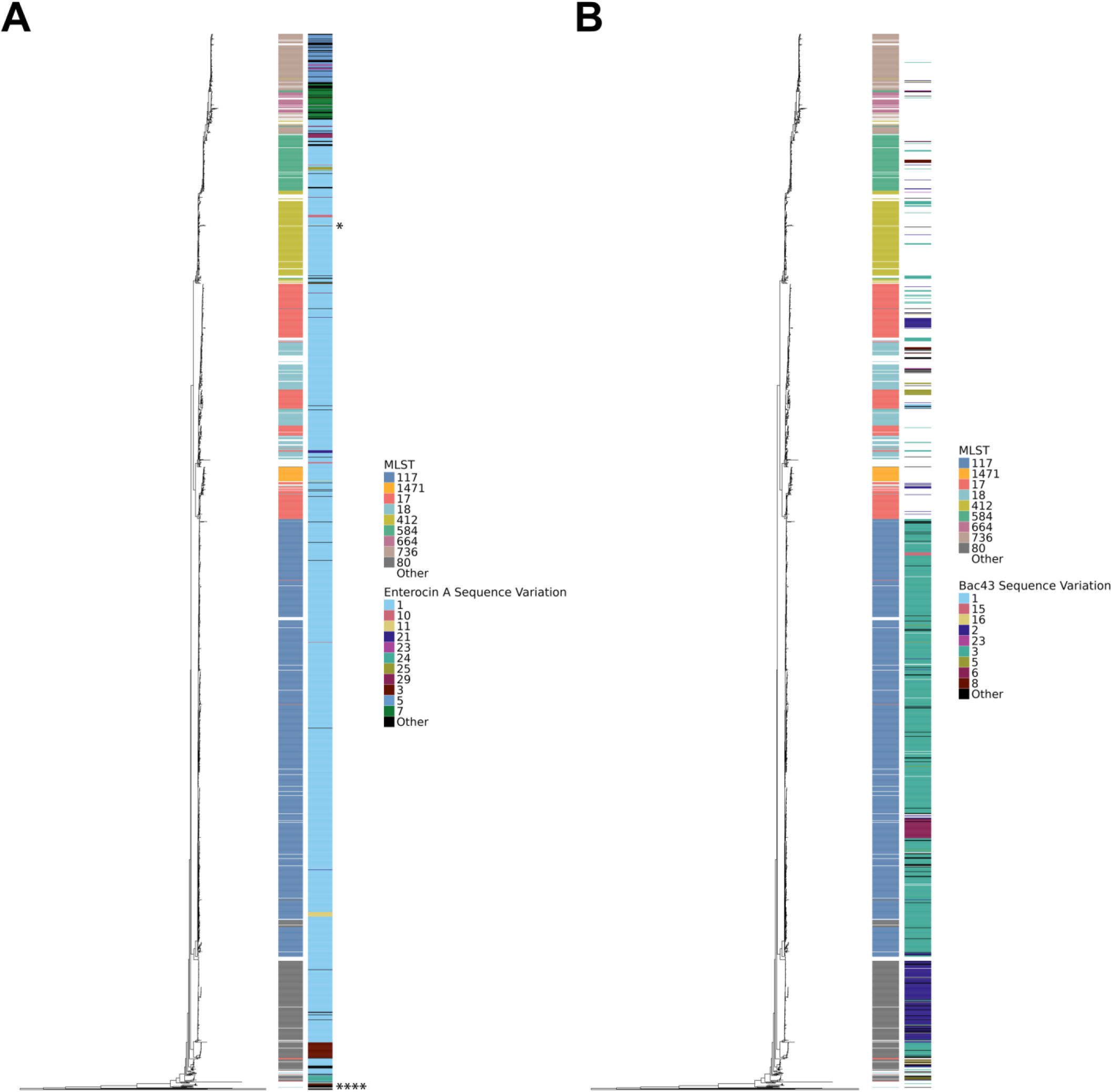
Phylogenetic mapping of bacteriocin genotype and phenotype. Core genome phylogeny with recombination removed by Gubbins. Outermost heatmap represents the bacteriocin sequence. Sequences with less than five isolates are classified as “Other.” Innermost heatmap is the isolate’s MLST. MLSTs with less than 20 isolates are classified as “Other.” A. Enterocin A sequences. Enterocin A isolates with the full enterocin A locus and an inhibitory phenotype are each indicated by an “*” B. Bac43 sequences.

For bac43, similar sequences tend to cluster together consistent with prominence of vertical transmission (Figure 7B). Sequence 3 is mostly found within MLST 117 and sequence 2 is associated with MLST 80. However, occasionally the same sequence can be found in distant clades supporting horizontal transmission events. Functional bac43 is found in 18 of the 59 MLSTs (seven of nine MLSTs found in more than 20 isolates).

### Temporal trends

The frequency of bac43 increased over the study period, rising from 8% in the first quarter of 2016 to 91% presence by the end of 2021 (Figure 8A). At the beginning of the collection period, no single MLST was present in more than 20% of isolates. However, MLST 117 subsequently rose to become the dominant sequence type with MLST 80 steadily increasing (Figure 8B). Isolates encoding bac43 are highly associated with MLSTs 117 and 80 accounting for 66.6% and 18.5% of bac43 isolates, respectively. By contrast, the six isolates that demonstrated functional enterocin A activity were dispersed over only the first five years of the study and had no clear association with MLSTs.

**Figure 8:**
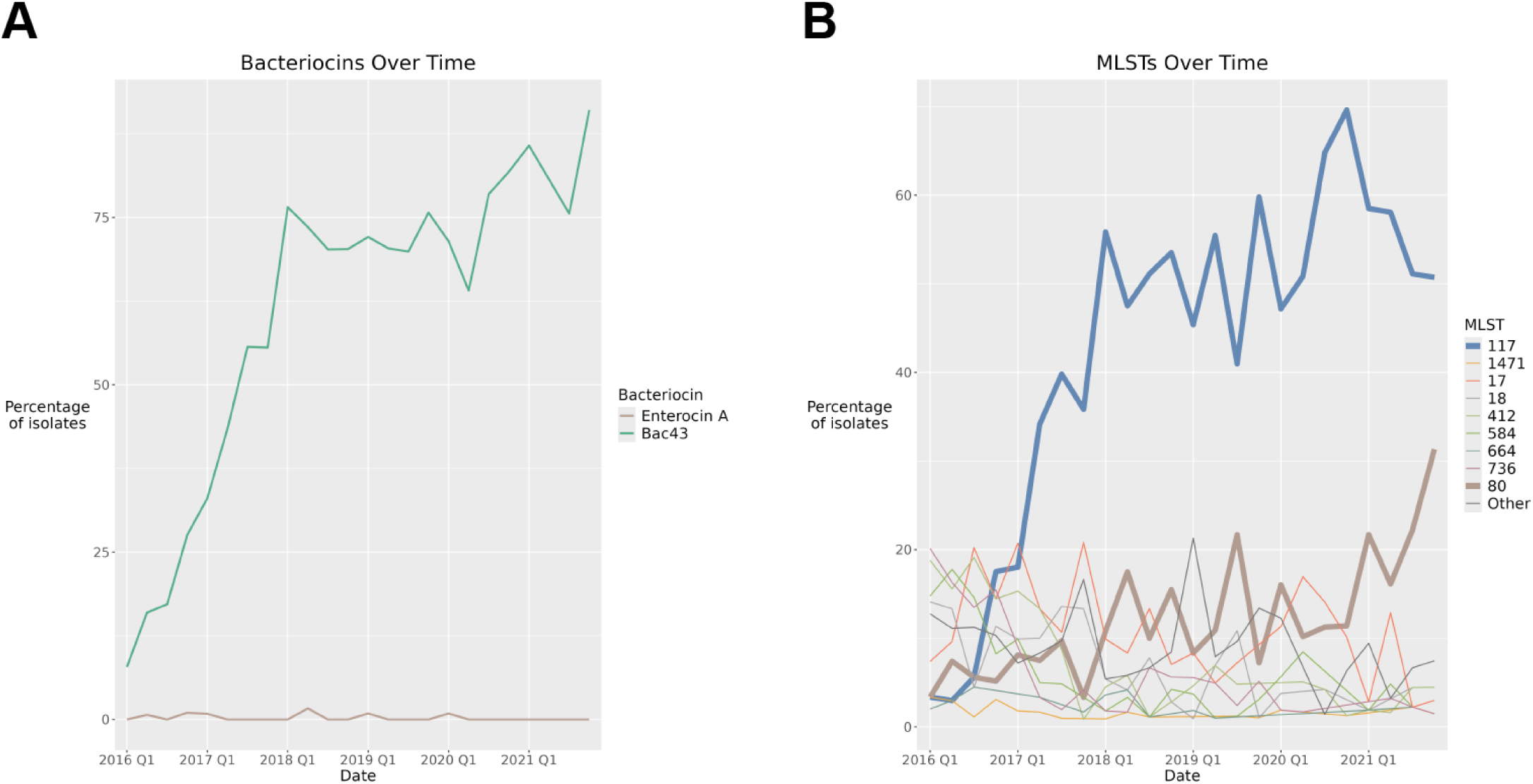
Temporal trends in bacteriocin variants and MLST. Presence of (A) functional bac43 and enterocin A sequences by quarter year and (B) MLSTs over the six-year study period, over the study period by quarter year. MLSTs with less than 20 isolates are classified as “Other.” Data includes first isolate per patient only.

### Prevalence by isolate source

Phenotypically functional enterocin A was found in 0.09% of perirectal surveillance swab (PR) isolates and 1.9% of blood (BL) isolates with significantly more enterocin A isolates in the BL cohort (p-value 0.00028)(Table 2). Conversely, bac43 is present in significantly more PR isolates (p-value 0.0086). However, our collection protocol only identified vancomycin resistant PR isolates, while BL includes both vancomycin resistant and vancomycin sensitive isolates. When we restrict the comparison to isolates that were vancomycin resistant, there is no significant differences in bacteriocin presence between the PR and BL isolates (Table 3).

**Table 2:**
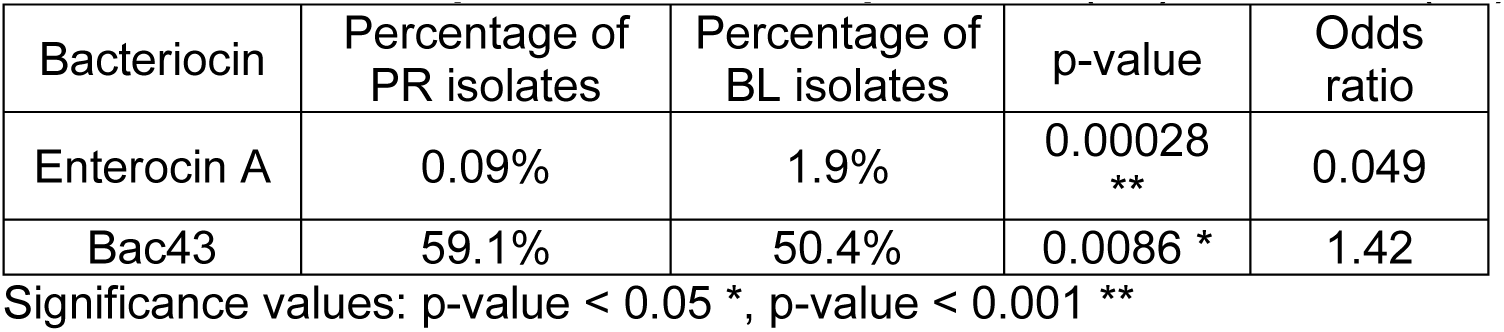
Bacteriocin presence between perirectal (PR) and blood (BL) isolates.

| Bacteriocin | Percentage of PR isolates | Percentage of BL isolates | p-value | Odds ratio |
| --- | --- | --- | --- | --- |
| Enterocin A | 0.09% | 1.9% | 0.00028<br>** | 0.049 |
| Bac43 | 59.1% | 50.4% | 0.0086 * | 1.42 |

Significance values: p-value < 0.05 \*, p-value < 0.001 \*\*

**Table 3:** Bacteriocin presence between vancomycin resistant perirectal (VRE PR) and vancomycin resistant blood (VRE BL) isolates.

| Bacteriocin | Percentage of VRE PR isolates | Percentage of VRE BL isolates | p-value | Odds ratio |
| --- | --- | --- | --- | --- |
| Enterocin A | 0.09% | 0% | 1 | Infinity |
| Bac43 | 59.1% | 56.9% | 0.62 | 1.09 |

Significance values: p-value < 0.05 \*, p-value < 0.001 \*\*

## Discussion

Enteroccocci occupy the intestinal tract of a broad array of host species and compete intensively with other inhabitants of this niche(5). Bacteriocins are commonly found in enterococci and believed to be effective weapons in this competition(6), but little is known about the role they play in clinical settings. Here we investigate the epidemiology and evolution of bacteriocins in *E. faecium* in a clinical setting by analyzing bacteriocin genetic content and phenotype of patient-derived isolates collected over a six-year period. Bacteriocin 43 (bac43) stands out as being the most common functional bacteriocin and shows a dramatic increase in frequency over time.

Bac43 was first isolated from a collection of VRE isolates from this same hospital in the late 1990s(30). From those isolates collected over five years, bac43 was identified in 3.3%. From our current study, we find the bac43 genes and its encoding plasmid to be mostly conserved with 40% of our isolates encoding a plasmid identical to the initially described plasmid(30). When considering both sets of data, we see that bac43 has been present within this hospital for over two decades with a recent rise in frequency. However, this observational genomic survey cannot identify a causal role for bac43 in this increasing frequency of over time. It is tempting to suggest bac43 could impact colonization and transmission given the strong phenotype it produces in the lab. Indeed, prior work using a mouse model of intestinal colonization showed that *E. faecalis* harboring a plasmid encoded bacteriocin bac-21 were able to eliminate bac-21 sensitive strains over the course of their experiment(10). That study also demonstrated bac-21 plasmid was actively conjugated within the GI tract(10). If pDT1 acts similarly in being advantageous to *E. faecium* for invasion and domination of the human gut, this could explain how bac43 has come to dominate our clinical cohort. However, there are alternative explanations. The strong association between bac43 and the MLSTs that have become dominant in our cohort (117 and 80), suggest these could be rising due to hitchhiking with some other features of these genomes. It is also possible that bac43 could be acting as a selfish plasmid, mostly serving to maintain its own existence with little benefit to the host bacterium. Finally, we have focused on the intraspecific competition, but it is also possible that bac43 could impact interspecific competition, especially among closely related species(30).

Prior studies have identified bac43 from clinical isolates of vancomycin resistant and sensitive enterococci (30, 46, 47) and demonstrated a correlation between bac43 and vancomycin resistant *E. faecium*(47). Consistently, we also find that bac43 is associated with vancomycin resistance. After correcting for this association, we see a similar presence of bac43 in strains from both colonization and infection.

This study also adds clarity to the contribution of enterocin A through genomic and phenotypic analyses. While the structural gene for enterocin A is found in 98% of *E. faecium* isolates within this cohort, the vast majority of these appear to be non-functional. Seven of the 11 sequences with a full enterocin A locus (0.3% of isolates) demonstrated an inhibitory phenotype. We also identified three isolates with the full enterocin A locus that did not have an inhibitory phenotype but did appear to be resistant to killing by enterocin A. While these isolates still retain the enterocin A immunity gene, this resistance could also reflect mutations in the bacteriocin target site, mannose phosphotransferase system(48).

Caution should be taken when interpreting the phenotypic results, as the inhibition and resistance assays we performed could be confounded by the presence of other bacteriocins. When present, other bacteriocins may result in killing of the sensitive strain used in the assay. Additionally, apparent resistance to the enterocin A producer could be due to other bacteriocin loci conferring resistance to enterocin A, or to suppression of the growth of the enterocin A producer. The presence of other bacteriocins may explain why several isolates that had only portions of the enterocin A locus and were not predicted to be functional had both inhibition and enterocin A immunity. Future work to elucidate the molecular function of the diverse bacteriocin loci described in this paper are warranted.

This study represents a systematic analyses of bacteriocins from a clinical cohort and identifies bacteriocin 43 (bac43) as being mostly conserved and increasing in prevalence in clinical *E. faecium* isolates over a six-year collection period. As bacteriocins have been shown to be effective weapons in bacterial competition, bac43 is an important target for future investigation of the role of bacteriocins in the transmission of vancomycin resistant *E. faecium*.

## Acknowledgements

AG was supported by the Molecular Mechanisms in Microbial Pathogenesis Training Program (T32 AI007528) and the University of Michigan Integrated Training in Microbial Systems (grant from Burroughs Wellcome Fund: Institutional Program Unifying Population and Laboratory Based Sciences). This work was supported by the National Institutes of Health under Grant No. R01 AI143852 to R.J.W.

## Supplemental data

**Supplemental Table 1:** Genes for enterocin A, bac43, and enterocin NKR-5-3B.

| Bacteriocin. | Gene | Length (bp) |
| --- | --- | --- |
| Enterocin A<br>(Genbank accession number AF099088.1) | entA | 198 |
|  | entI | 312 |
|  | entF | 147 |
|  | entK | 1284 |
|  | entR | 753 |
|  | entT | 2154 |
|  | entD | 1368 |
| Bac43<br>(Genbank accession number AB178871.1) | bacA | 225 |
|  | bacB | 288 |
|  | mobC | 381 |
|  | mobA | 915 |
|  | repA | 945 |
|  | repB | 534 |
| Enterocin NKR-5-3B<br>(Genbank accession number LC068607.1) | enkB | 264 |
|  | enkB1 | 1140 |
|  | enkB2 | 519 |
|  | enkB3 | 591 |
|  | enkB4 | 498 |

**Supplemental Table 2:**
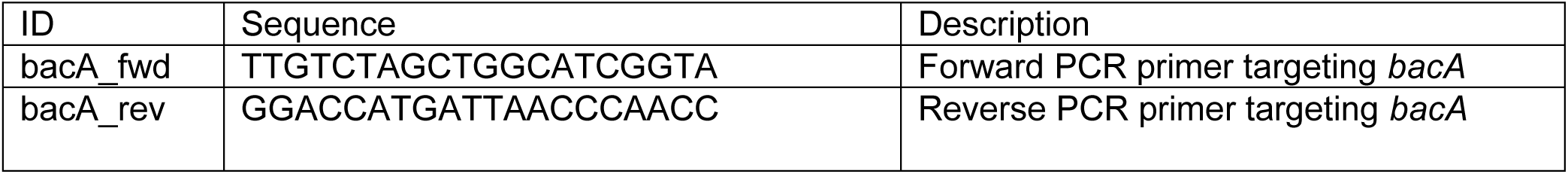
PCR primers used for confirming bacteriocin gene presence.

**Supplemental Table 3:** All bacteriocin cluster hits and the number of isolates they are found in.

| **Bacteriocin cluster** | **Isolate presence** |
| --- | --- |
| Enterocin A cluster | 2395 |
| Bac43 full cluster | 1406 |
| Bac43 partial cluster | 988 |
| Enterocin NKR-5-3B cluster | 466 |
| Enterocin SE-K4 cluster | 69 |
| Enterocin P cluster | 14 |
| Enterocin B cluster | 13 |
| Bac32 cluster | 12 |
| Enterocin Xα cluster | 10 |
| Bac32 truncated cluster | 5 |
| Enterocin Xβ cluster | 4 |
| Enterocin L50b cluster | 3 |
| Enterocin L50a cluster | 3 |
| Enterocin Q cluster | 2 |
| Pneumolancidin cluster | 1 |
| Bac31 cluster | 1 |
| Enterocin P-like cluster | 1 |

